# Feasibility and Sensitivity of Saliva GeneXpert MTB/RIF Ultra for Tuberculosis Diagnosis in Adults in Uganda

**DOI:** 10.1101/2022.03.16.22272031

**Authors:** Patrick Byanyima, Sylvia Kaswabuli, Emmanuel Musisi, Catherine Nabakiibi, Josephine Zawedde, Ingvar Sanyu, Abdul Sessolo, Alfred Andama, William Worodria, Laurence Huang, J. Lucian Davis

## Abstract

The objective of this prospective, observational study carried out at China-Uganda Friendship Hospital-Naguru in Kampala, Uganda, was to determine the performance of GeneXpert MTB/RIF Ultra (Xpert) testing on saliva for active tuberculosis (TB) disease among consecutive adults undergoing diagnostic evaluation. We calculated sensitivity to determine the diagnostic performance in comparison to that of the composite reference standard of *Mycobacterium tuberculosis* liquid and solid cultures on two spot sputum specimens. GeneXpert Ultra on saliva had a sensitivity of 90% (95% confidence interval [CI], 81-96%); this was similar to that of sputum fluorescence smear microscopy (FM) of 87% (95% CI, 77-94%). Sensitivity was 24% lower (95% CI for difference 2-48%, p=0.003) among persons living with HIV (71%, 95%CI 44-90%) than among persons living without HIV (95%, 95%CI 86-99%) and 46% lower (95% CI for difference 14-77%, p<0.0001) among sputum microscopy positive (96%, 95% CI 87-99%) than among sputum microscopy negative patients (50%, 95% CI 19-81%). Semi-quantitative Xpert grade was higher in sputum than in paired saliva samples from the same patient. In conclusion, saliva specimens appear to be feasible and similarly sensitive to sputum for active TB diagnosis using molecular testing, suggesting promise as a non-sputum diagnostic test for active TB in high-burden settings.

## INTRODUCTION

**O**ver the last quarter century, improvements in diagnosis and treatment of people with tuberculosis (TB) have gradually reduced mortality, but large gaps in detection and treatment persist that contribute to substantial ongoing morbidity and mortality [1]. Among several available strategies to facilitate rapid, same-day diagnosis of TB, testing sputum with the GeneXpert MTB/RIF Ultra molecular assay [2, 3] is the most sensitive and most readily available approach. Unfortunately, there are several operational challenges associated with collecting sputum for diagnosis of pulmonary TB. First, coughing during sputum expectoration or sputum induction generates aerosols that may facilitate TB transmission [4]. Second, some individuals are unable to produce sputum, including young children, those with dry cough, and the severely ill/severely debilitated. Given these limitations of sputum for TB diagnosis, in 2014 the World Health Organization (WHO) issued guidelines for developers of a future non-sputum test for active TB diagnosis, including a target product profile suggesting that it should have a minimum diagnostic accuracy similar to sputum GeneXpert MTB/RIF on sputum smear-negative individuals (*i*.*e*., sensitivity ≥68%, specificity ≥98%) [5].

One alternative sample type with great promise for diagnosis of pulmonary TB is saliva, which is easy to collect, with minimal risk of generating aerosols. Although Stop TB Partnership guidelines discourage collection of salivary sputum samples because they have lower diagnostic yield for acid-fast bacilli (AFB) by microscopy or culture, the diagnostic yield of TB molecular testing appears to be more promising. In a previous study of 1782 smear-negative adults undergoing evaluation for active TB, for example, we found that salivary sputum provided a substantially higher diagnostic yield and sensitivity for culture-positive TB than other sputum types, implying incremental value to using oral samples at least as a supplement to sputum [6]. Using a different sampling technique, oral swabs, Wood and colleagues showed that oral nylon swabs repeatedly tested positive for TB via IS6110 polymerase chain reaction testing in 90% of South African patients with TB confirmed by sputum GeneXpert MTB/RIF testing, suggesting that TB is present in the oral cavity [7]. A subsequent study of 50 adults with possible TB in Uganda found similar sensitivity of 88%, albeit with lower specificity. Saliva is also now widely used for molecular diagnosis of COVID-19, where it has high sensitivity, even among patients without symptoms [8]. Using saliva as a diagnostic specimen in the COVID-19 context has been shown to reduce aerosol exposure for health workers and eliminate the need for personal protective equipment because it is self-collected [9]. This raises the possibility that saliva alone could be used as a TB diagnostic when paired with next generation and ultra-sensitive molecular tests (GeneXpert MTB/RIF Ultra). Thus, the aim of this study was to evaluate the feasibility and sensitivity of GeneXpert MTB/RIF on saliva among symptomatic adult TB confirmed patients.

## MATERIALS AND METHODS

### Study design & Population

Between June 2018 and May 2019, we carried out a prospective, observational study to determine the performance of GeneXpert MTB/RIF Ultra (Xpert) testing on saliva for diagnosis of active TB. This was a sub-study nested within the Mulago Inpatient Non-invasive Diagnosis of Pneumonia–Inflammation Aging, Microbes, and Obstructive Lung Disease (I AM OLD) study. We enrolled consecutive adults (age ≥18 years) with cough of any duration but *<*6 months who were also undergoing TB evaluation (including HIV testing, chest radiography, and sputum examination) as inpatients or outpatients at China-Uganda Friendship Hospital-Naguru in Kampala, Uganda; patients with a prior history of TB within the past two years and those receiving treatment for active TB at the time of presentation were excluded. In this sub-study, we included consecutive patients with positive sputum Xpert results at any semi-quantitative threshold.

### Measurements and Study Procedures

After obtaining written informed consent from participants, a study nurse collected demographic and clinical information using a structured questionnaire, and then provided standardized instructions to expectorate sputum into three separate cups “on the spot” [10]. Trained study staff examined the first sample using direct auramine-O fluorescence microscopy (FM) [11, 12] and sent the remaining sample for mycobacterial culture and speciaion on Lowenstein-Jensen (LJ) solid media and in Mycobacterial Growth Indicator Tube (MGIT) liquid media, the accepted microbiologic reference standard assays for TB. Staff examined the second sample using direct FM and performed GeneXpert MTB/RIF testing on the remainder [13]. Finally, staff sent a third sputum sample for mycobacterial culture on solid media and liquid culture.. All cultures were performed at the Makerere University Mycobacteriology Laboratory, and staff performing the cultures were not provided with clinical information about the study participants. At least two hours after sputum collection, the patients were asked to submit at least 1 mL of saliva placed into a sterile specimen cup for GeneXpert MTB/RIF testing; all participants were instructed not to cough prior to saliva collection. Saliva specimens were processed for GeneXpert MTB/RIF using a sample reagent to saliva volume ratio of 1:1, and all other steps followed the manufacturer’s recommendations for extra-pulmonary body fluid specimens [13]. Sputum was collected prior to TB treatment initiation, and saliva was collected prior to or within two hours of TB treatment initiation. Finally, all participants without a prior known HIV diagnosis were offered HIV testing and counseling, and for those found to be living with HIV, a CD4+ T-cell count was performed at the Makerere University–Johns Hopkins University Research Collaboration (MU-JHU) laboratory.

### Statistical Analysis

We examined baseline characteristics using proportions for categorical variables, and medians for continuous variables. We calculated sensitivity for GeneXpert MTB/RIF results on saliva and on sputum in reference to a composite reference standard described as follows: those with ≥1 sputum sample culture-positive were defined as *Mycobacterium tuberculosis (Mtb)* positive, those with two negative cultures were defined as negative, and all others were defined as indeterminate. We estimated precision using exact binomial 95% confidence intervals. We explored comparisons of diagnostic accuracy results (sensitivity differences with 95% CI) and semi-quantitative results (Fisher’s exact test) for saliva GeneXpert by sputum smear microscopy and HIV status. We estimated that a sample size of 84 patients would enable us to determine if the sensitivity of saliva GeneXpert MTB/RIF was 75% or higher with a precision of ±10%, allowing for up 10% indeterminate results due to missing or contaminated sputum culture results. We used STATA 14.0 (Stata Corporation, College Station, TX) for all statistical analyses.

### Human subjects protection

The study protocol was reviewed and approved by the Yale University and the University of California San Francisco Institutional Review Boards, the Makerere University School of Medicine Research Ethics Committee, the Mulago Hospital Institutional Review Board, and the Uganda National Council for Science and Technology.

#### Data sharing

A comprehensive, de-identified dataset containing individual-level data will be made available prior to publication.

## RESULTS

### Study Population

Among 153 participants enrolled into the parent study between June 2018 and June 2019, 40 were GeneXpert MTB/RIF negative; 15 did not have sputum GeneXpert MTB/RIF performed; 16 did not provide a saliva specimen; and one participant had an indeterminate culture result, (Figure 1) leaving 81 participants for inclusion in the analysis. There were no adverse events during specimen collection. Median age of participants was 30 years (interquartile range 24-38), 50 (62%) were men. 18 (22%) were persons living with HIV, with median CD4 cell count 90 cells/uL (interquartile range 49-234), and only seven of the 18 (39%) were taking antiretroviral therapy at enrolment. 17 (21%) had ever smoked ≥100 cigarettes in their entire life and 60 (74%) had a history of alcohol use. 17 (21%) had a cough greater than two weeks, while 71 (88%) reported subjective fever within the past seven days. 75 (93%) reported weight loss, including 46 (57%) with weight loss ≥5 kg. 14 (17%) reported no ambulatory limitation; 47 (58%) were mildly limited with ambulation, and 20 (25%) were severely affected but not bedbound. 67 patients (83%) were AFB smear-positive, including 30 (37%) with an AFB microscopy smear grade of 3+, 18 (22%) with a grade of 2+, nine (11%) 1+, and 10 (12%) had 1-9 AFB seen per 100 high-powered fields. 13 (16%) were AFB smear-negative and one (1%) was missing an AFB smear microscopy result (Table 1).

**TABLE 1.**
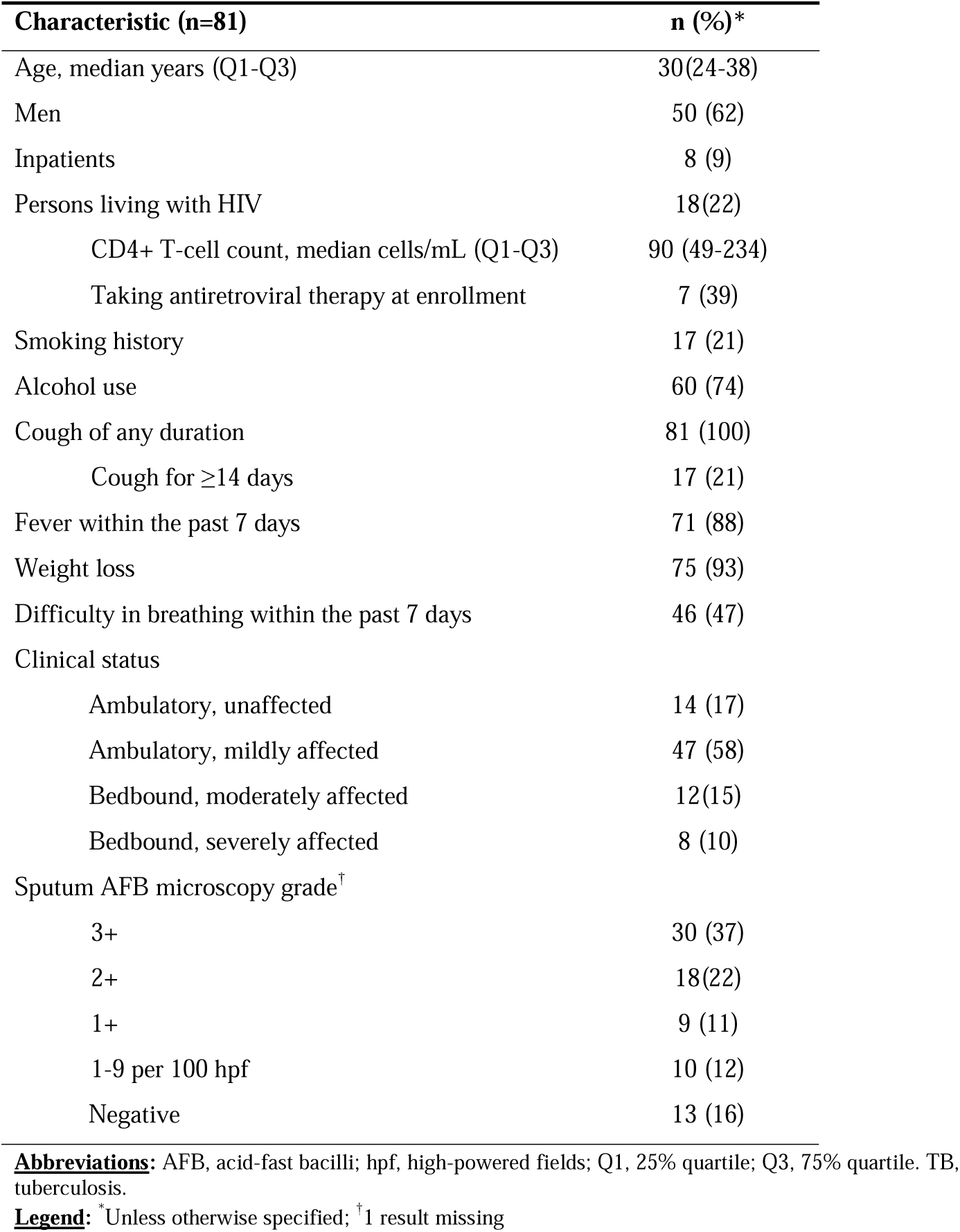
Demographic and clinical characteristics.

| **Characteristic (n=81)** | **n (%)\*** |
| --- | --- |
| Age, median years (Q1-Q3) | 30(24-38) |
| Men | 50 (62) |
| Inpatients | 8 (9) |
| Persons living with HIV | 18(22) |
| CD4+ T-cell count, median cells/mL (Q1-Q3) | 90 (49-234) |
| Taking antiretroviral therapy at enrollment | 7 (39) |
| Smoking history | 17 (21) |
| Alcohol use | 60 (74) |
| Cough of any duration | 81 (100) |
| Cough for $\geq 14$ days | 17 (21) |
| Fever within the past 7 days | 71 (88) |
| Weight loss | 75 (93) |
| Difficulty in breathing within the past 7 days | 46 (47) |
| Clinical status |  |
| Ambulatory, unaffected | 14 (17) |
| Ambulatory, mildly affected | 47 (58) |
| Bedbound, moderately affected | 12(15) |
| Bedbound, severely affected | 8 (10) |
| Sputum AFB microscopy grade<sup>†</sup> |  |
| 3+ | 30 (37) |
| 2+ | 18(22) |
| 1+ | 9 (11) |
| 1-9 per 100 hpf | 10 (12) |
| Negative | 13 (16) |

**Abbreviations:** AFB, acid-fast bacilli; hpf, high-powered fields; Q1, 25% quartile; Q3, 75% quartile. TB, tuberculosis.
**Legend:** \* Unless otherwise specified; <sup>†</sup> 1 result missing

**FIGURE 1.**
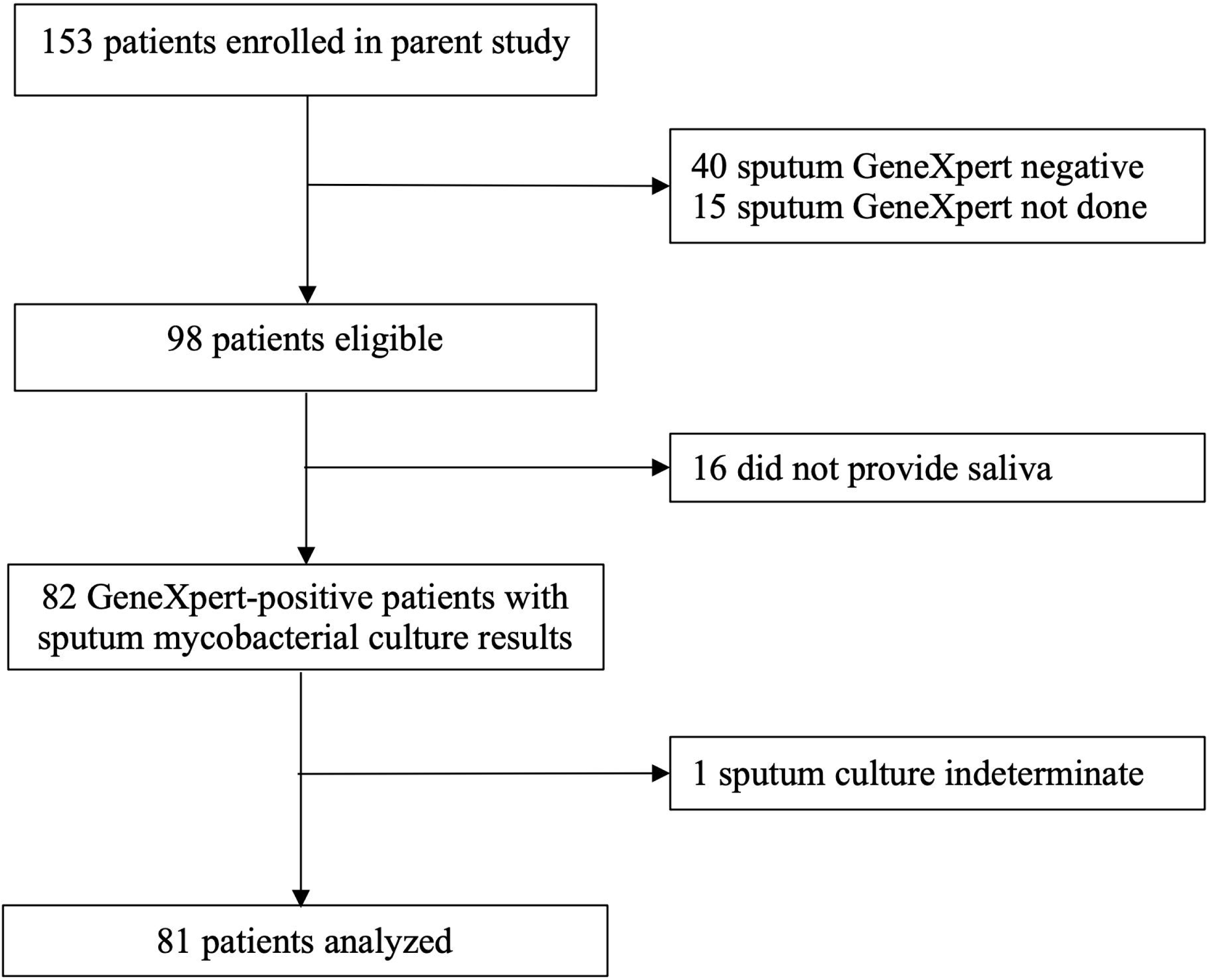
Flow diagram showing study enrollment and TB testing results. Patients missing index test results due to a missing saliva sample and patients with missing reference standard results due indeterminate culture results were excluded from analysis.

### Diagnostic Performance

Seventy-eight patients were confirmed *Mtb* culture-positive on liquid and/or solid media, while three were *Mtb* culture-negative. Seventy-three of the 78 patients with culture-confirmed TB were salivary GeneXpert MTB/RIF positive, giving an overall sensitivity of GeneXpert MTB/RIF on saliva of 90% (95% Confidence Interval (CI) 81-96%). This sensitivity was similar to that of sputum smear microscopy, which had a sensitivity of 87% (95% CI 77-94%) in reference to the combined culture reference standard. Among the three *Mtb* culture-negative patients, two were salivary GeneXpert positive. Sensitivity was 24% lower (95% CI for difference 2-48%, p=0.003) among persons living with HIV (71%, 95% CI 44-90%) than among persons living without HIV (95%, 95% CI 86-99%), and 46% lower (95% CI for difference 14-77%, p<0.0001) among sputum microscopy positive (96%, 95% CI 87-99%) than among sputum microscopy negative patients (50%, 95% CI 19-81%).

We also compared the semi-quantitative results of bacilli by GeneXpert for both saliva and sputum, as shown in Table 2. Overall, the semi-quantitative GeneXpert grade was higher in sputum samples than in paired saliva samples collected from the same patient: 56 of 72 (78%) of the sputum samples of either medium (n=22) or high (n=34) semi-quantitative grade, whereas only 14 of 72 (19%) of the saliva samples were of either medium (n=10) or high (n=4) grade, indicating that the mycobacterial load in the saliva specimens was low overall. There was no difference in semi-quantitative results by smear microscopy result (p=0.52) or by HIV status (p=0.39).

**TABLE 2.** Within-individual comparisons of semi-quantitative GeneXpert MTB/RIF Ultra results between sputum and saliva, among those with positive test results on both sample types (n=72).

| **Sputum**<br>**Xpert**<br>**Results** | **Saliva Xpert Results** |  |  |  |  |
| --- | --- | --- | --- | --- | --- |
|  | Trace | Very Low | Low | Moderate | High |
| Trace | 0 | 0 | 0 | 0 | 0 |
| Very Low | 0 | 0 | 1 | 0 | 0 |
| Low | 2 | 5 | 6 | 0 | 2 |
| Moderate | 0 | 4 | 15 | 2 | 1 |
| High | 2 | 3 | 20 | 8 | 1 |

**Legend:** Shading intensity is proportional to the frequency of paired results by semi-quantitative grade across the two sample types.

## DISCUSSION

In a prospective, observational study of consecutive sputum GeneXpert-positive TB patients in a high-burden setting, we showed that diagnosis of TB using GeneXpert Ultra on saliva is feasible and had a high sensitivity relative to a rigorously defined reference standard. This finding has significant implications for the diagnosis of TB and potentially also for TB infection control. Using sputum specimens for TB diagnosis poses a number of challenges, since some individuals such as those with non-productive cough and young children find expectoration challenging, and the associated generation of sputum aerosols poses an infection control risk for health care workers and nearby patients [14]. The development of novel testing strategies that employ non-sputum samples for TB has been identified as a priority by the WHO, and the sensitivity measured in our study is consistent with WHO’s minimum target-product profile for a non-sputum-based test, with similar sensitivity to sputum GeneXpert among a population of predominantly sputum microscopy-positive and HIV-negative individuals. Although our alternative strategy of salivary GeneXpert exceeds WHO’s optimal targets for cost ($4) and turn-around time (20 minutes) for a non-sputum-based test, if GeneXpert on saliva were shown to perform well in populations for whom sputum collection is less feasible for the reasons described above, the willingness to pay for and wait for results might be higher.

The use of saliva for molecular diagnosis of TB was first described in a convenience sample of 52 adult TB patients in Japan who were evaluated using a lab-developed, nested PCR assay that was shown to have a sensitivity of 98% [15]. A more recent study of 44 sputum smear- and culture-positive TB patients, including 35 in South Africa and 9 in South Korea, reported on saliva as having a very low sensitivity of 39% for TB testing [16]. Sputum mycobacterial load was similarly high (100% smear-positive in the South Africa/South Korea study vs. 87% in our study), so these differences in diagnostic performance might be attributable to differences in either sample collection or specimen processing. For example, participants were instructed to rinse their mouths prior to specimen collection in the South Africa/South Korea study but not in our study. Second, the South Africa/South Korea study diluted one part of the sample in two parts of sample reagent as recommended by the manufacturer for sputum, while we used a 1:1 dilution ratio as recommended for cerebrospinal fluid, another extra-pulmonary specimen without a mucoid matrix [17]. Finally, we used the GeneXpert MTB/RIF Ultra cartridge, which has ten-fold better analytic sensitivity than the earlier generation GeneXpert MTB/RIF cartridge. To our knowledge, we are among the first to report the performance of GeneXpert MTB/RIF Ultra on saliva.

Previous studies have examined the sensitivity of a variety or oral specimens for diagnosis of TB. We previously showed that oropharyngeal wash specimens paired with a lab-developed PCR assay had a high sensitivity for TB diagnosis in reference to sputum mycobacterial culture on previously frozen and thawed sputum, but a subsequent study failed to confirm these results [18, 19]. A study of *Mtb* PCR on buccal swabs of South African TB patients and US controls showed high sensitivity (90%) and specificity (100%), although the case-control design may have inflated diagnostic accuracy [7]. A recent study from the US was among the first to show that saliva is a viable and accurate specimen for diagnosis of SARS-CoV2, and more sensitive and less variable than nasopharyngeal swab specimens [20]. Another study carried out in Thailand using saliva for diagnosis of SARS-CoV2 showed similar results, with saliva providing a sensitivity of 84% and a specificity of 99% [21]. Collectively, these studies suggest that saliva is a very promising novel specimen for diagnosis of respiratory tract infections.

There were a few limitations to our study. First, because the primary study objective was to evaluate feasibility and preliminary sensitivity, we did not include patients with non-productive cough or children, two ideal target populations for salivary testing. If, as seems plausible, these populations have more paucibacillary disease, diagnostic sensitivity could be lower in these populations, as suggested by the lower sensitivity observed among sputum smear-negative individuals and persons living with HIV. However, in the current study, we found that even though saliva is more paucibacillary than sputum as assessed by GeneXpert’s semi-quantitative measurement of mycobacterial load, diagnostic sensitivity was similar between the two specimen types, likely because of the extremely low threshold of analytic sensitivity of the GeneXpert Ultra assay [22]. Secondly, to conserve costs in this preliminary study, we did not enroll non-TB patients to serve as controls, a choice that prevented us from estimating diagnostic specificity. However, a recent systematic review found that both GeneXpert MTB/RIF assays have a high specificity on a variety of body fluid types [23]. Thirdly, our sample size was small, especially for persons living with HIV and for sputum smear-negative patients, which limited our ability to develop precise accuracy estimates for these and other subgroups.

In conclusion, saliva appears to be a feasible specimen for TB diagnosis using GeneXpert Ultra, with a similar diagnostic sensitivity to sputum GeneXpert Ultra, at least among HIV-negative and sputum smear-positive individuals. and thus appear to be a very promising alternative non-sputum diagnostic test for active TB in high-burden settings. Future studies should examine sensitivity in populations who are most likely to benefit from this test, including individuals who are unable to expectorate sputum, children, and individuals from populations with a broad spectrum of mycobacterial load and disease severity, and symptomatic individuals without TB, including persons living with HIV. Direct comparisons of saliva to other oral sampling methods, including swabs, would also be useful. Finally, studies evaluating the relative impacts of salivary versus sputum testing on infection control proxies and/or on outcomes would also be valuable.

## Supporting information

STARD Checklist

## Data Availability

A comprehensive, de-identified dataset containing individual-level data will be made available prior to publication online at the Dryad website.

## ACKNOWLEDGMENTS

The authors would like to acknowledge the patients who participated in the study and the administration and TB clinic staff of the China-Uganda Friendship Hospital where the study took place.

